# Polygenic and *Polygene x Trauma* Contributions to Alcohol Use and Problems Among Black Americans

**DOI:** 10.1101/2025.05.13.25327255

**Authors:** Chelsie E. Benca-Bachman, Samantha Cassidy, Rameez A. Syed, Whitney L. Barfield, Natalia Jaume-Feliciosi, Alicia K. Smith, Seyma Katrinli, Abigail Powers, Yara Mekawi, Rohan H. C. Palmer

## Abstract

**Aims:** To estimate polygenic and polygene x environment contributions to alcohol consumption and problems in the context of childhood maltreatment and lifetime trauma.

**Design:** Main and interaction effects models predicting alcohol consumption and problems were estimated using multiple linear regression. Covariates included age, sex, education, employment status, and ancestral principal components.

**Setting:** USA

**Participants:** A sample of 2,114 Black adults (75% female; M_age_=39.88, SD=13.92) recruited from the Grady Trauma Hospital in Atlanta, Georgia.

**Measurement:** Polygenic scores (PGS) for trauma-related symptoms (re-experiencing: PGS_REEX_, avoidance: PGS_AVOID_, hyperarousal: PGS_HYPER_, PTSD symptom score: PGS_PCL_) and alcohol consumption (PGS_AUDIT-C_) and use disorder (AUD; PGS_AUD_) were derived using genome-wide association Million Veterans Program summary statistics with PRS-CS.

**Findings:** Childhood maltreatment and lifetime trauma (excluding childhood abuse) were positively associated with alcohol consumption (AUDIT-C) (β_childhood-maltreatment_=0.17, SE=0.02; β_lifetime-trauma_=0.28, SE=0.02) and alcohol problems (AUDIT-P) (β_childhood-maltreatment_=0.25, SE=0.02; β_lifetime-trauma_=0.27, SE=0.02). None of the PGSs were associated with AUDIT-C, but both the PGSs for re-experiencing (β=0.1, SE=0.03) and avoidance (β=0.08, SE=0.03) were positively associated with AUDIT-P. Experiencing lifetime trauma and being at elevated genetic risk for AUD (interaction-β_Trauma_x_PGSAUD_=0.17, SE=0.05) and hyperarousal (interaction-β_TraumaxPGSHYPER_=0.11, SE=0.06) were associated with higher AUDIT-P scores; while more lifetime trauma and higher genetic risk for AUD were associated with higher AUDIT-C scores (interaction-β_Trauma_x_PGSAUD_=0.12, SE=0.05).

**Conclusions:** Individuals with elevated genetic risk for AUD are more likely to consume alcohol and to develop worse alcohol problems in the context of lifetime trauma. Interventions focused on also minimizing the effects of trauma-exposure would be particularly beneficial among individuals at risk for AUD.

## INTRODUCTION

Alcohol is the most used addictive substance in the United States (1) and its misuse is associated with significant morbidity, mortality, disability, and deterioration of quality of life (2). Using data from the 2023 National Survey on Drug Use and Health, the 12-month point prevalence of alcohol use disorder in people aged 18 and older was 10.9% (3). Due to the health burden that problematic alcohol use poses, it is crucial to understand the environmental and genetic factors that influence individuals to increase alcohol consumption and problematic alcohol use.

It is well established that some individuals may start or escalate alcohol use (or other substance use) as a way to cope with trauma (4). Trauma refers to incidents of physical, emotional, or life-threatening harm that impacts an individual’s mental, physical, and/or emotional health, as well as social or spiritual well-being (5). Traumatic events can be natural disasters, accidents, or interpersonal violence. Generally, prior traumatic experiences during childhood and adulthood have been linked to increased alcohol consumption and problems generally (6, 7). Additionally, there are documented longitudinal relationships between trauma and alcohol misuse. For instance, research by Berenz and colleagues (8) found that interpersonal trauma was associated with increased drinking around the time of the trauma and afterwards for college-aged women, but not men. Furthermore, experiencing trauma is associated with partaking in risky behaviors, which include substance use (9). Overall, there is growing evidence that drinking to cope in the face of trauma may mediate observations between problematic drinking and trauma- and stress-related disorders (10). However, the relationship between trauma and alcohol use is not unidirectional. Alcohol and other substance use has been linked to behaviors and scenarios that may increase the likelihood of experiencing a traumatic event. For example, emergency room department data suggests that 10-18% of accidental injuries are attributable to alcohol consumption (11). Additionally, alcohol and substance use has been linked to increased aggression and violence (12). Preexisting alcohol use may also be a risk factor for experiencing more post-traumatic stress disorder (PTSD) symptoms after a traumatic event. Tripp and colleagues (13) found that not only did PTSD symptoms predict future alcohol use, but more alcohol consumption predicted the future severity of PTSD symptoms. Taken together, these findings illustrate that people who consume greater quantities of alcohol may be more likely to experience events that are traumatic and that can lead to PTSD.

While not everyone who experiences trauma develops PTSD, most people experience at least one traumatic event in their lifetime (14). Kilpatrick and colleagues (2013) found that 89.7% of adults had experienced at least one traumatic event, and the mode number of traumatic events participants reported experiencing was three. People who experience interpersonal violence traumas, particularly sexual violence, are more likely to develop PTSD compared to other types of traumas (15). There are also gender differences in trauma exposure - women are more likely to report sexual violence and childhood abuse whereas men are more likely to report nonsexual violence, living through a natural disaster, and being in an accident (16). Schein and colleagues (2021) found in their systematic review that the 12-month point prevalence of PTSD within civilians ranges from 2.3-9.1%, and that the military personnel 12-month point prevalence has a large range from 6.7-50.2% (17). Women (16) and Black individuals are also more likely to develop PTSD (18), making it particularly important to examine associations in this population. People who engage in maladaptive coping mechanisms and report high levels of neuroticism are more likely to experience PTSD symptoms (19). In addition to trauma and drinking, genetics is another risk factor for PTSD. Sartor and colleagues (20) examined the heritable influences on PTSD and found that genetic variation contributed to 46% of the variance in liability. Additionally, Stein and colleagues (2021) conducted a genome-wide association study (GWAS) on three of the PTSD criteria: hyperarousal, re-experiencing, and avoidance; total PTSD Checklist for DSM-IV (PCL) was also included in the GWAS (21). Numerous genetic loci were identified to be associated with each criterion examined and the PCL, suggesting that these phenotypes are polygenic. Of those who experienced trauma and developed PTSD, 40% had comorbid alcohol use disorder (AUD) (22). This is not to imply that having AUD is necessary or sufficient for developing PTSD. According to the Diagnostic and Statistical Manual of Mental Disorders (DSM-5), in order to be diagnosed with PTSD individuals need to (A) be exposed to a stressor (i.e., a traumatic event) and in response to that stressor endorse experiencing a number of symptoms that fall within four criterion domains: (B) intrusion, (C) avoidance, (D) negative alterations in cognitions and mood, and (E) alterations in arousal and reactivity (23). While most research has looked at PTSD diagnosis or symptom severity in relation to alcohol use, some studies have found associations with PTSD symptom subdomains. More symptoms of hyperarousal, negative alterations in cognition and mood, avoidance, and re-experiencing were all associated with greater problematic alcohol consumption (24, 25). The relationship between the two also appears bidirectional; as alcohol consumption has also been shown to predict greater likelihood of developing PTSD symptoms in the future (13). Indeed, recent evidence indicates that there is also a genetic association between AUD and PTSD in both twin (*r*_*g*_=.54 (26)) and population studies (*r*_*g*_=.35 (21)). Furthermore, a recent genomic structural equational modeling study found that both AUD and alcohol use are genetically correlated with PTSD (broadly defined; *r*_*g*_= .36 and *r*_*g*_=−0.17, respectively), and that different genetic variants might contribute to these two relationships (i.e., alcohol use and AUD with PTSD) (27). Similarly, Palmer and colleagues (28) used genome-wide effects to identify a partial genetic correlation (*r*_*g*_=.49) between experiencing interpersonal trauma and vulnerability to drug dependence.

The current study aimed to better understand the extent to which common polygenic variation related to alcohol use, AUD, and PTSD phenotypes, in the form of polygenic scores (PGSs), explains individual differences in alcohol problems in Black individuals who have previously experienced trauma. Based on the diathesis stress model we hypothesized that (1) childhood maltreatment or prior traumatic experiences (excluding childhood abuse) would be associated with increased alcohol problems and alcohol consumption. Further (2), individuals with predisposing genetic liability for alcohol use, AUD, or PTSD would evidence even greater alcohol use and severity of disorder.

## METHODS

### Participants

#### GWAS discovery data

Participants for the summary statistics used in this study were of African ancestry Veterans from the Million Veterans Program (MVP; dbGaP accession phs001672.v6.p1 (29)). Overall, the MVP data is approximately 77% white, 13% Black, and 90% male. The exact N’s and percentages varied depending on the phenotype and when in the data collection process each GWAS was conducted. Summary statistics for six phenotypes were used in the current set of analyses to generate PGSs: alcohol consumption (PGS_AUDIT-C_), AUD, PTSD checklist total score (PGS_PCL)_, and PTSD sub-scores for reexperiencing (PGS_REEX_), avoidance (PGS_AVOID_), and hyperarousal (PGS_HYPER_). Given that cross-ancestry PGS prediction has generally been low (30, 31), we used summary statistics from the African ancestry cohorts given our target dataset was comprised of African ancestry individuals. Table 1 and Supplementary Methods show the number of participants and summary statistics for the study measures described below.

**Table 1.** Number of Participants and Variants Contributing to GWAS Summary Statistics.

| Phenotype | AFR Sample N | AFR Variant K | Overlap with HapMap K | EUR Sample N | EUR Variant K | Citations |
| --- | --- | --- | --- | --- | --- | --- |
|  | African Ancestry |  |  | European Ancestry |  |  |
| AUD | 56,648 | 12,542,083 | 419,069 | 202,004 | 6,895,251 | Kranzler et al., 2019 |
| AUDIT-C | 56,495 | 12,545,885 | 419,418 | 200,680 | 6,898,150 | Kranzler et al., 2019 |
| HYPER | 25,521 | 15,727,245 | 1,183,468 | 186,689 | 9,388,996 | Stein et al., 2021 |
| REEX | 25,521 | 15,726,271 | 1,183,462 | 186,689 | 9,369,491 | Stein et al., 2021 |
| PCL | 25,414 | 16,339,113 | 1,201,282 | 186,689 | 9,369,975 | Stein et al., 2021 |
| AVOID | 25,414 | 15,726,047 | 1,183,464 | 186,689 | 9,388,940 | Stein et al., 2021 |

*Note.* N=number of participants; K = number of genetic variants; AUD = Alcohol Use Disorder; AUDIT-C = AUDIT consumption subscore; HYPER= Hyperarousal; REEX= Reexperiencing; PCL = PCL total score; AVOID= Avoidance. Overlap with HapMap K indicates the number of variants from the summary statistics that overlap with HapMap3 reference panel variants used for the PGS methods.

#### Target data

Target data came from the Grady Trauma Project (GTP). Procedures for data collection are outlined in Mekawi and colleagues (2020) (32). A total of 5,248 self-identified African American participants provided genetic and phenotypic information, with a smaller subset of 2,114 individuals who provided information on their drinking behaviors which were used for the current analyses. The current study incorporated analyses for two outcome measures: alcohol consumption and alcohol problems. Participant ages ranged between 18 and 82 years (*M*_age_ = 39.89 years old, *SD* = 13.17). Individuals in our sample predominantly self-identified as female (75%); See Table 2 for descriptive information. The current secondary data analysis study was approved by the Emory University Institutional Review Board (IRB00090295).

**Table 2.** Descriptive Statistics.

| Variable | N | Mean/% | SD | Median | Min | Max | Skew | Kurtosis | SE |
| --- | --- | --- | --- | --- | --- | --- | --- | --- | --- |
| Sex (female) | 2124 | 73% | N/A | N/A | N/A | N/A | N/A | N/A | N/A |
| Age | 2117 | 39.89 | 13.17 | 41 | 18 | 78 | 0.03 | -1.17 | 0.29 |
| Age<sup>2</sup> | 2117 | 1764.45 | 1067.45 | 1681 | 324 | 6084 | 0.45 | -0.71 | 23.2 |
| Education | 2112 | 1.87 | 1.62 | 1 | 0 | 6 | 0.64 | -0.56 | 0.04 |
| Employment Status (employed) | 2110 | 31% | N/A | N/A | N/A | N/A | N/A | N/A | N/A |
| Income | 2054 | 2.09 | 1.37 | 2 | 0 | 4 | -0.27 | -1.13 | 0.03 |
| TEI | 2103 | 3.98 | 2.65 | 4 | 0 | 14 | 0.64 | 0.08 | 0.06 |
| CTQ | 2100 | 40.73 | 17.93 | 34 | 25 | 120 | 1.6 | 2.13 | 0.39 |
| AUDIT-C | 2114 | 5.26 | 3.85 | 4 | 0 | 12 | 0.38 | -1.17 | 0.08 |
| AUDIT-P | 2086 | 4.31 | 6.4 | 1 | 0 | 28 | 1.59 | 1.59 | 0.14 |

*Note.* SD = Standard Deviation; Min = Minimum; Max = Maximum; SE = Standard Error; N/A = Not Applicable; TEI = Trauma Events Inventory; CTQ = Childhood Trauma Questionnaire.

### Assessments

#### Demographics

Age was assessed with the question “How old are you?”. Participants were asked “What is your sex?” and response options were male or female (coded as 0 and 1 respectively). Education was assessed with the question, “What was the highest grade you completed in school?”.

Responses were re-coded to the following categories: less than high school, completed high school or obtained a GED, or more than high school. Income was assessed with the question, “What is your approximate household monthly income?” and responses were recoded into categories (0,1, or 2): less than $500 per month, less than $1000 per month, or more than $999 per month. Employment status was assessed with the question, “Are you currently employed?”, and response options were, “No” (coded as 0) or “Yes” (1). When participants chose to not answer a given question the responses were coded as missing.

#### Childhood maltreatment

Childhood maltreatment was assessed using the Childhood Trauma Questionnaire (CTQ) (33). Participants were instructed to answer based on their childhood and adolescence and responded with a Likert scale where 1 corresponded with “Never True” and 5 corresponded with “Always True”. Three of the 28 items (“There was nothing I wanted to change about my family”, “I had the perfect childhood”, “I had the best family in the world” decreased the measure’s overall Cronbach’s alpha and so those items were removed. A raw total score was created using the mean across the items, which was then multiplied by 25. This approach allowed us to calculate scores for anyone who was missing up to 3 items. The resulting Cronbach’s alpha was .94.

#### Traumatic experiences

Lifetime trauma was assessed through the Traumatic Events Inventory (TEI), which included 15 items (34). This self-report instrument measures lifetime (childhood and adulthood) exposure to trauma such as natural disaster, serious accident or injury, and physical or sexual assault, as well as frequency of events, age at worst incident, and feelings of terror, horror, and helplessness. The TEI, which was developed for use with the GTP study population. For the current study, only items that inquired about situations participants self-report either experiencing or witnessing d throughout the lifetime were included (as opposed to situations the participant had heard of happening to someone close to them). Items that measured maltreatment during childhood were excluded to reduce overlap with the CTQ. Participants endorsed the occurrence (no or yes coded as 0 or 1 respectively). Raw TEI scores were operationalized as a continuous measure, ranging from 0 to 14. The observed Cronbach’s alpha was .71.

#### Alcohol consumption

Alcohol consumption was assessed using the first three items of the Alcohol Use Disorders Identification Test (AUDIT) (35). These items asked about alcohol consumption during the year the participant drank the most. An example of an item was, “During the year when you drank the most, how often did you have a drink containing alcohol?”. Participants responded to each item using a Likert scale, ranging from 0 (“Never”) to 4 (“4 or more times a week”). The items were summed together to create a composite score. The maximum raw score possible was 12.

#### Alcohol problems

Alcohol problems were assessed using the remaining seven items of the AUDIT (35). These items asked about alcohol problems during the year the participant drank the most. An example of an item was, “During the year you drank the most, how often did you find that you were unable to stop drinking once you started?”. Items were summed together to create a composite score. The maximum raw score possible was 28.

### Genotyping, quality control, and imputation of target data

Global ancestry was first determined, and subjects categorized as African American were imputed and analyzed separately (36). Data were imputed based on the 1000 Genomes phase 3 data (1KGP phase 3)(37). Details on global ancestry determination, quality control, and imputation can be found in supplemental information and prior publications (38).

### Polygenic score derivation

PGSs were created using PRS-CS (39) using the African ancestry summary statistics referenced in Table 1. PRS-CS utilizes continuous shrinkage priors to adjust GWAS summary statistics weights while accounting for Linkage Disequilibrium (LD) to derive polygenic score weights. We used precomputed LD matrices from the African population in 1000 Genomes Phase 3 as our LD reference panel (https://github.com/getian107/PRScs). PGSs were generated using only the variants that matched with the HapMap3 reference panel (40), resulting in approximately 1.3 million overlapping SNP. We ran PRS-CS using all autosomal chromosomes. For the GWAS sample size parameter, each study’s total participant N (see Table 1) was used rather than varying N by SNP. Input summary statistics datasets were formatted, and default parameters were used in PRS-CS. Lastly, we used the PLINK [version 1.9] (41) score function to apply weights to the target GTP dataset. Additionally, PGSs were calculated using PRS-CSX utilizing the African ancestry summary statistics in conjunction with the European ancestry summary statistics to see if there was a boost in signal due to increased summary statistic sample sizes (42). Findings with the multi-ancestry PRS-CSX scores were largely consistent with the African ancestry PRS-CS scores. As such, the African Ancestry results are presented using PRS-CS scores; PRS-CSX results are available upon request.

### Ancestral principal components

To control for allele frequency variation due to admixture, principal components (PCs) were created using the smartPCA algorithm in Eigenstrat. Participants that were classified as African ancestry via global ancestry determination were included in analyses. The first five PCs were included as covariates in analyses to control for residual population stratification.

### Analytic strategy

All variables underwent descriptive screening using R for Statistics (43). Right-skewed variables for the CTQ, TEI, alcohol consumption, and alcohol problems were log-transformed after adding one to each score to better approximate normality. These log-transformed measures were included in analyses.

#### Determination of main and interaction effects of PGSs and trauma phenotypes on alcohol outcomes

To assess the main and interaction effects of the PGSs and the trauma phenotypes on the alcohol outcome variables we used Mplus (version 8) (44). First, we calculated the descriptive statistics and Cronbach’s alpha when appropriate. Second, we examined the zero-order correlations between the variables. Third, we ran a series of increasingly complex regression models that examined the effects of each PGS and type of trauma/maltreatment on the alcohol outcomes plus demographic covariates and genetic ancestry PCs. The first series of regression model equations were:

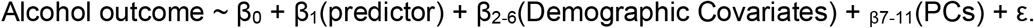

We assessed each of the main effects of the PGSs on the alcohol outcomes in the context of trauma by including each PGS individually in the model and the two trauma phenotypes as the predictors and repeating this with each PGS. Model 2’s regression equation was the following:

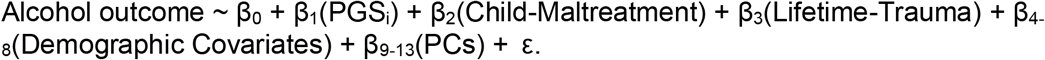

We also assessed the interaction effects of each PGS by looking at the moderation between each PGS and the maltreatment/trauma outcomes while in the context of trauma. The regression equation for the moderation models were as follows:

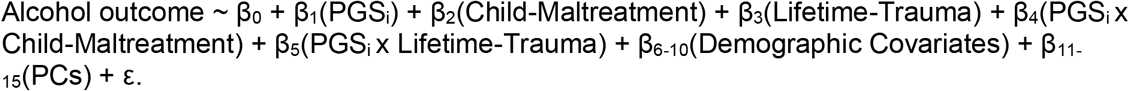

Finally, when there were significant PGS effects, we ran a joint model that included all the PGSs together while controlling for the effects of trauma to understand if the PGS explained unique variation in the alcohol outcomes or if they were potentially explaining shared variance.

## RESULTS

### Descriptive statistics & correlations

Descriptive statistics are available in Table 2. Briefly, 40.3% of the overall sample provided information related to their alcohol consumption (*N*=2114) and alcohol problems (N=2086). AUDIT-C sub-scores suggested low-moderate use on average (*M*=5.26; *SD*=3.85). Similarly, AUDIT-P sub-scores indicated that participants experienced a moderate level of problems (*M*=4.31; *SD*=6.40). Zero-order correlations in Table 3 describe the relationships between study variables. Lifetime trauma and childhood maltreatment were moderately correlated (*r* = .40), such that experiencing more maltreatment during childhood was associated with reporting more overall trauma exposure. Most demographic covariates (e.g., sex and education level), were also associated with both types of traumas. Interestingly, these covariates showed small to moderate correlations with the alcohol PGSs (r’s ranged from -.08 to -.43), but only education was negatively associated with the PTSD PGSs (r’s were consistently - .07). Our two outcomes, AUDIT-C and AUDIT-P were also positively correlated (*r* =.70), such that those who had more severe problems tended to consume more alcohol. The PGS_PCL_ was positively correlated with the PTSD subdomain PGSs (*r*’s ranged from .90 to.94). Despite these high correlations, the PTSD-PGSs showed differential relationships with many variables, while the alcohol PGSs (PGSAUD and PGS_AUDIT-C_) had small, but significant correlations with PGS_PCL_ and the other PTSD-PGSs (*r*’s .11 to .17). Lifetime trauma was significantly related to the PTSD PGSs (*r*’s -.03 to -.04), but not the alcohol PGSs. The only PGS that childhood maltreatment was significantly correlated with was the PGS_AUDIT-C_ (*r* = .03).

**Table 3.** Zero-order correlations between study variables.

|  |  | Descriptive Phenos |  |  |  |  | PTSD PGSSs |  |  |  | Alcohol PGSSs |  | Trauma Phenos |  | Alcohol Phenos |  |
| --- | --- | --- | --- | --- | --- | --- | --- | --- | --- | --- | --- | --- | --- | --- | --- | --- |
|  |  | Sex | Age | Edu | Empl Status | Income | PCL | REEX | HYPER | AVOID | AUDIT-C | AUD | TEI Log | CTQ Log | AUDIT-C Log | AUDIT-P Log |
| Descriptive Phenos | Sex | 1.0 |  |  |  |  |  |  |  |  |  |  |  |  |  |  |
|  | Age | **-0.27†** | 1.0 |  |  |  |  |  |  |  |  |  |  |  |  |  |
|  | Education | **0.06\*\*** | **0.03\*** | 1.0 |  |  |  |  |  |  |  |  |  |  |  |  |
|  | Empl Status | **0.11†** | **-0.22†** | **0.23†** | 1.0 |  |  |  |  |  |  |  |  |  |  |  |
|  | Income | **0.16†** | -0.00 | **0.20†** | **0.50†** | 1.0 |  |  |  |  |  |  |  |  |  |  |
| PTSD PGSSs | PCL | 0.03 | -0.01 | **-0.07†** | -0.01 | 0.00 | 1.0 |  |  |  |  |  |  |  |  |  |
|  | Reex | 0.02 | -0.00 | **-0.07†** | -0.02 | -0.01 | **0.94†** | 1.0 |  |  |  |  |  |  |  |  |
|  | Hyper | 0.02 | -0.01 | **-0.07†** | -0.01 | -0.00 | **0.90†** | **0.84†** | 1.0 |  |  |  |  |  |  |  |
|  | Avoid | 0.03 | -0.00 | **-0.07†** | -0.01 | -0.01 | **0.94†** | **0.91†** | **0.84†** | 1.0 |  |  |  |  |  |  |
| Alcohol PGSSs | AUDIT-C | 0.02 | -0.00 | -0.03 | 0.03 | 0.00 | **0.13†** | **0.13†** | **0.11†** | **0.13†** | 1.0 |  |  |  |  |  |
|  | AUD | 0.01 | 0.01 | -0.02 | 0.02 | -0.02 | **0.15†** | **0.15†** | **0.12†** | **0.17†** | **0.55†** | 1.0 |  |  |  |  |
| Trauma Phenos | TEI Log | **-0.26†** | **0.20†** | **0.08†** | **-0.09†** | -0.01 | -<br>**0.04\*\*** | **-0.03\*** | **-0.03\*** | -<br>**0.04\*\*** | -0.01 | 0.01 | 1.0 |  |  |  |
|  | CTQ Log | **0.12†** | -0.00 | **-0.03\*** | **-0.08†** | **-0.09†** | -0.02 | -0.02 | -0.02 | -0.01 | **0.03\*** | 0.02 | **0.40†** | 1.0 |  |  |
| Alcohol Phenos | AUDIT-C | **-0.43†** | **0.28†** | -<br>**0.08\*\*** | **-0.11†** | **-0.09\*\*** | 0.02 | 0.03 | 0.02 | 0.01 | -0.01 | 0.01 | **0.35†** | **0.12†** | 1.0 |  |
|  | AUDIT-P | **-0.36†** | **0.20†** | **-0.17†** | **-0.14†** | **-0.17†** | 0.03 | **0.05\*** | 0.03 | 0.04 | 0.03 | 0.04 | **0.32†** | **0.21†** | **0.70†** | 1.0 |

*Note.* Bolded values met a $p < .05$ threshold. Phenos = Phenotypes; PGS = Polygenic score; Edu = Education; Empl = Employment; TEI = Trauma Events Inventory; CTQ = Childhood Trauma Questionnaire; AUDIT-C = Alcohol Use Disorders Identification Test; AUD = Alcohol Use Disorder; REEX = Reexperiencing PTSD subdomain; HYPER = Hyperarousal PTSD subdomain; AVOID = Avoidance PTSD subdomain; PCL= PTSD Checklist Total score; Log = log transformed variable.

**Table 4.** Polygenic Effects on Alcohol Consumption while Controlling for Trauma.

| Model | 1. PGS<sub>AUDIT-C</sub> | 2. PGS<sub>AUD</sub> | 3. PGS<sub>REEX</sub> | 4. PGS<sub>HYPER</sub> | 5. PGS<sub>AVOID</sub> | 6. PGS<sub>PCL</sub> |
| --- | --- | --- | --- | --- | --- | --- |
|  | beta (se) | beta (se) | beta (se) | beta (se) | beta (se) | beta (se) |
| PGS | 0.01 (0.02) | 0.02 (0.02) | 0.02 (0.03) | 0.02 (0.02) | 0.01 (0.03) | 0.01 (0.03) |
| TEI log | **0.25 (0.02)** | **0.25 (0.02)** | **0.25 (0.02)** | **0.25 (0.02)** | **0.25 (0.02)** | **0.25 (0.02)** |
| CTQ log | **0.06 (0.02)** | **0.06 (0.02)** | **0.06 (0.02)** | **0.06 (0.02)** | **0.06 (0.02)** | **0.06 (0.02)** |
| Sex | **-0.23 (0.02)** | **-0.23 (0.02)** | **-0.23 (0.02)** | **-0.23 (0.02)** | **-0.23 (0.02)** | **-0.23 (0.02)** |
| Age | **0.19 (0.02)** | **0.19 (0.02)** | **0.19 (0.02)** | **0.19 (0.02)** | **0.19 (0.02)** | **0.19 (0.02)** |
| Education | **-0.06 (0.02)** | **-0.06 (0.02)** | **-0.06 (0.02)** | **-0.06 (0.02)** | **-0.06 (0.02)** | **-0.06 (0.02)** |
| Employment | -0.02 (0.02) | -0.02 (0.02) | -0.02 (0.02) | -0.02 (0.02) | -0.02 (0.02) | -0.02 (0.02) |
| Income | -0.01 (0.02) | -0.01 (0.02) | -0.01 (0.02) | -0.01 (0.02) | -0.01 (0.02) | -0.01 (0.02) |
| PC1 | -0.02 (0.02) | -0.02 (0.02) | -0.01 (0.03) | -0.01 (0.02) | -0.01 (0.03) | -0.01 (0.03) |
| PC2 | 0.04 (0.02) | 0.04 (0.02) | 0.04 (0.02) | 0.04 (0.02) | 0.04 (0.02) | 0.04 (0.02) |
| PC3 | 0.02 (0.02) | 0.02 (0.02) | 0.02 (0.02) | 0.03 (0.02) | 0.02 (0.02) | 0.02 (0.02) |
| PC4 | 0.01 (0.02) | 0.01 (0.02) | 0.01 (0.02) | 0.01 (0.02) | 0.01 (0.02) | 0.01 (0.02) |
| PC5 | 0.03 (0.02) | 0.03 (0.02) | 0.03 (0.02) | 0.03 (0.02) | 0.03 (0.02) | 0.03 (0.02) |

*Note.* Each model (3-8) was run separately for a given PGS; More explicitly, model 3 included the AUDIT-C PGS as well as lifetime trauma, childhood maltreatment, demographic covariates and genetic ancestry PCs, whereas model 4 included the PGS for AUD as well as lifetime trauma, childhood maltreatment, demographic covariates and genetic ancestry PCs, etc. Bolded values met a $p < .05$ threshold; beta = beta estimate; se = standard error; PGS = Polygenic score; TEI = Trauma Events Inventory; CTQ = Childhood Trauma Questionnaire; AUDIT-C = Alcohol Use Disorders Identification Test; AUD = Alcohol Use Disorder; REEX = Reexperiencing PTSD subdomain; HYPER = Hyperarousal PTSD subdomain; AVOID = Avoidance PTSD subdomain; PCL= PTSD Checklist Total score; PC = Principal Component.

**Table 5.** Polygenic Effects on Alcohol Problems while Controlling for Trauma.

| Model | 1. PGS<sub>AUDIT-C</sub> | 2. PGS<sub>AUD</sub> | 3. PGS<sub>REEX</sub> | 4. PGS<sub>HYPER</sub> | 5. PGS<sub>AVOID</sub> | 6. PGS<sub>PCL</sub> |
| --- | --- | --- | --- | --- | --- | --- |
|  | beta (se) | beta (se) | beta (se) | beta (se) | beta (se) | beta (se) |
| PGS | 0.04 (0.02) | 0.03 (0.02) | **0.08 (0.03)** | 0.04 (0.02) | **0.07 (0.03)** | 0.04 (0.03) |
| TEI log | **0.20 (0.02)** | **0.20 (0.02)** | **0.20 (0.02)** | **0.20 (0.02)** | **0.20 (0.02)** | **0.20 (0.02)** |
| CTQ log | **0.16 (0.02)** | **0.16 (0.02)** | **0.16 (0.02)** | **0.16 (0.02)** | **0.16 (0.02)** | **0.16 (0.02)** |
| Sex | **-0.24 (0.02)** | **-0.24 (0.02)** | **-0.24 (0.02)** | **-0.24 (0.02)** | **-0.24 (0.02)** | **-0.24 (0.02)** |
| Age | **0.11 (0.02)** | **0.11 (0.02)** | **0.11 (0.02)** | **0.11 (0.02)** | **0.11 (0.02)** | **0.11 (0.02)** |
| Education | **-0.12 (0.02)** | **-0.12 (0.02)** | **-0.12 (0.02)** | **-0.12 (0.02)** | **-0.12 (0.02)** | **-0.12 (0.02)** |
| Employment | -0.01 (0.02) | -0.01 (0.02) | -0.01 (0.02) | -0.01 (0.02) | -0.01 (0.02) | -0.01 (0.02) |
| Income | **-0.07 (0.02)** | **-0.07 (0.02)** | **-0.07 (0.02)** | **-0.07 (0.02)** | **-0.07 (0.02)** | **-0.07 (0.02)** |
| PC1 | -0.01 (0.02) | -0.01 (0.02) | 0.04 (0.03) | 0.01 (0.02) | 0.03 (0.027) | 0.02 (0.03) |
| PC2 | 0.02 (0.02) | 0.02 (0.02) | 0.02 (0.02) | 0.02 (0.02) | 0.02 (0.02) | 0.02 (0.02) |
| PC3 | 0.03 (0.02) | 0.03 (0.02) | 0.03 (0.02) | 0.03 (0.02) | 0.04 (0.02) | 0.03 (0.02) |
| PC4 | 0.03 (0.02) | 0.03 (0.02) | **0.04 (0.02)** | 0.04 (0.02) | 0.04 (0.02) | 0.04 (0.02) |
| PC5 | 0.00 (0.02) | 0.00 (0.02) | 0.00 (0.02) | 0.00 (0.02) | -0.00 (0.02) | 0.00 (0.02) |

*Note.* Each model (3-8) was run separately for a given PGS as explained in the note for Table 2, but here the outcome is alcohol problems. Bolded values met a $p < .05$ threshold; beta = beta estimate; se = standard error; PGS = polygenic score; TEI = Trauma Events Inventory; CTQ = Childhood Trauma Questionnaire; AUDIT-C = Alcohol Use Disorders Identification Test; AUD = Alcohol Use Disorder; REEX= Reexperiencing PTSD subdomain; HYPER= Hyperarousal PTSD subdomain; AVOID= Avoidance PTSD subdomain; PCL= PTSD Checklist Total score; PC = Principal Component.

### Polygenic and gene x trauma effects on alcohol consumption and problems

Analytic findings of the effects of maltreatment and trauma and each PGS on alcohol consumption and use disorder, ignoring trauma history, are summarized in Supplementary Tables S1 and S2. While no PGS was associated with consumption, higher scores on all the PGSs were associated with having more alcohol problems (β estimates ranging from 0.05-0.27). Notably, several of these PGS effects were inflated due to the exclusion of history of childhood maltreatment and lifetime trauma in Model 1. Findings in the main effects model (Table S2), which controlled for history of childhood maltreatment and lifetime trauma, indicated that higher scores on the re-experiencing (*β*_REEX_=0.08; SE=0.03) and avoidance (*β*_REEX_=0.07; SE=0.03) PGSs were associated with greater alcohol problems. Similarly, greater childhood maltreatment and lifetime trauma were associated with increased alcohol consumption (Consumption: *β*_CTQ_=0.06 (SE=0.02) and *β*_TEI_=0.25 (0.02), respectively) and problems (Consumption: *β*_CTQ_=0.16 (SE=0.02) and *β*_TEI_=0.20 (0.02), respectively. Further expansion of the models to test our interaction hypotheses (Model 3) indicated that individuals who experienced trauma within their lifetime and had higher genetic risk for AUD consumed more alcohol on average (*β*_PGS:AUDxTEI_=0.12, SE=0.05; see Figure 1). Similarly, polygenic effects of AUD and hyperarousal on alcohol problems were exacerbated if individuals also experienced trauma within their lifetime ((*β*_PGS:AUDxTEI_=0.17, SE=0.05; Figure 2) and (*β*_PGS:HYPER x TEI_ =0.11, SE=0.06; Figure 3)). In other words, individuals who experienced a higher level of trauma within their lifetime and who also inherited a greater genetic liability for manifesting AUD or re-experiencing trauma presented with more alcohol problems.

**Figure 1.**
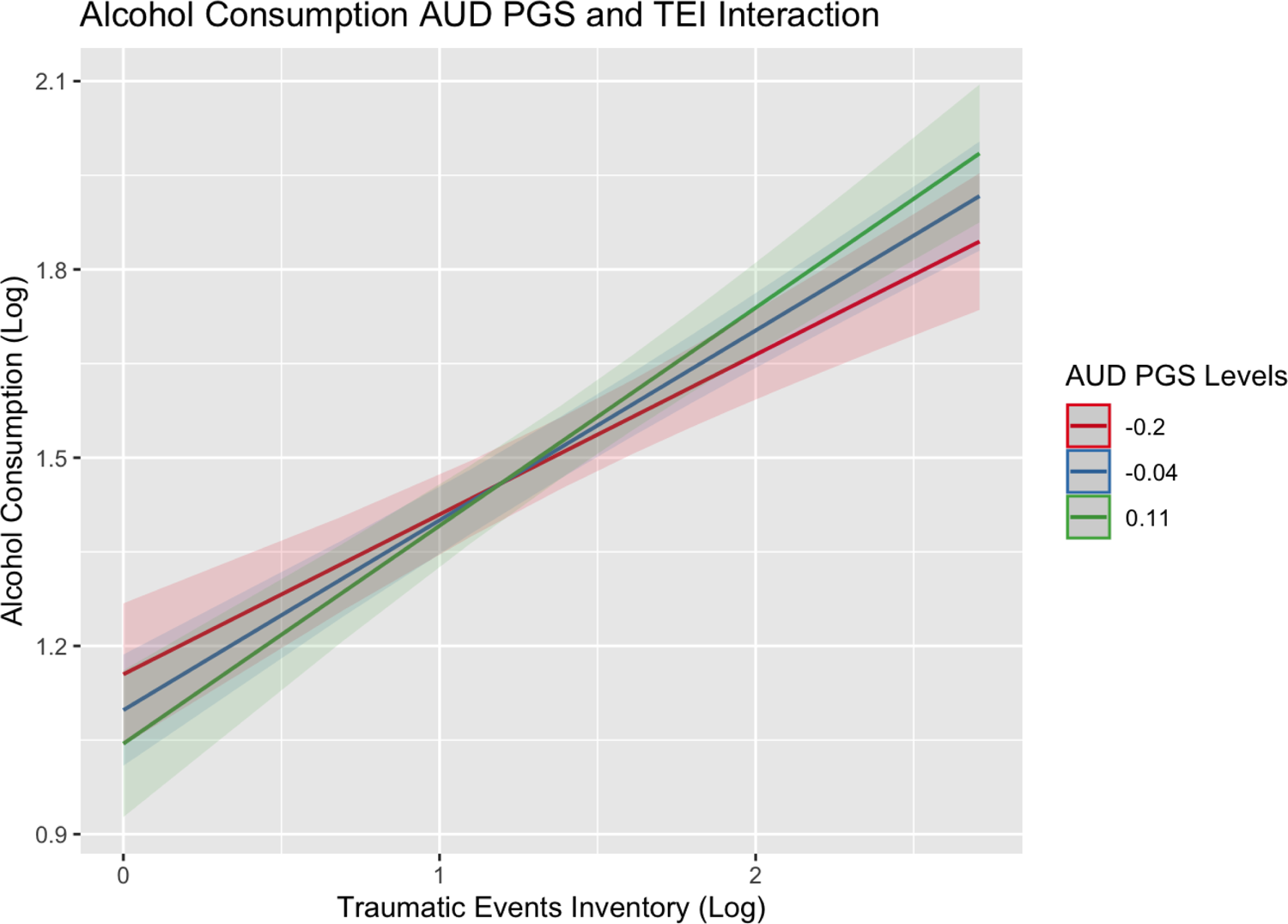
Alcohol consumption interaction plot for PGS_AUD_ and TEI Figure showing the effects of lifetime trauma exposure on levels of alcohol consumption among individuals with a PGS_AUD_ score two standard deviations below the mean (red), at the mean (blue), or above the mean (green) of the sample.

**Figure 2.**
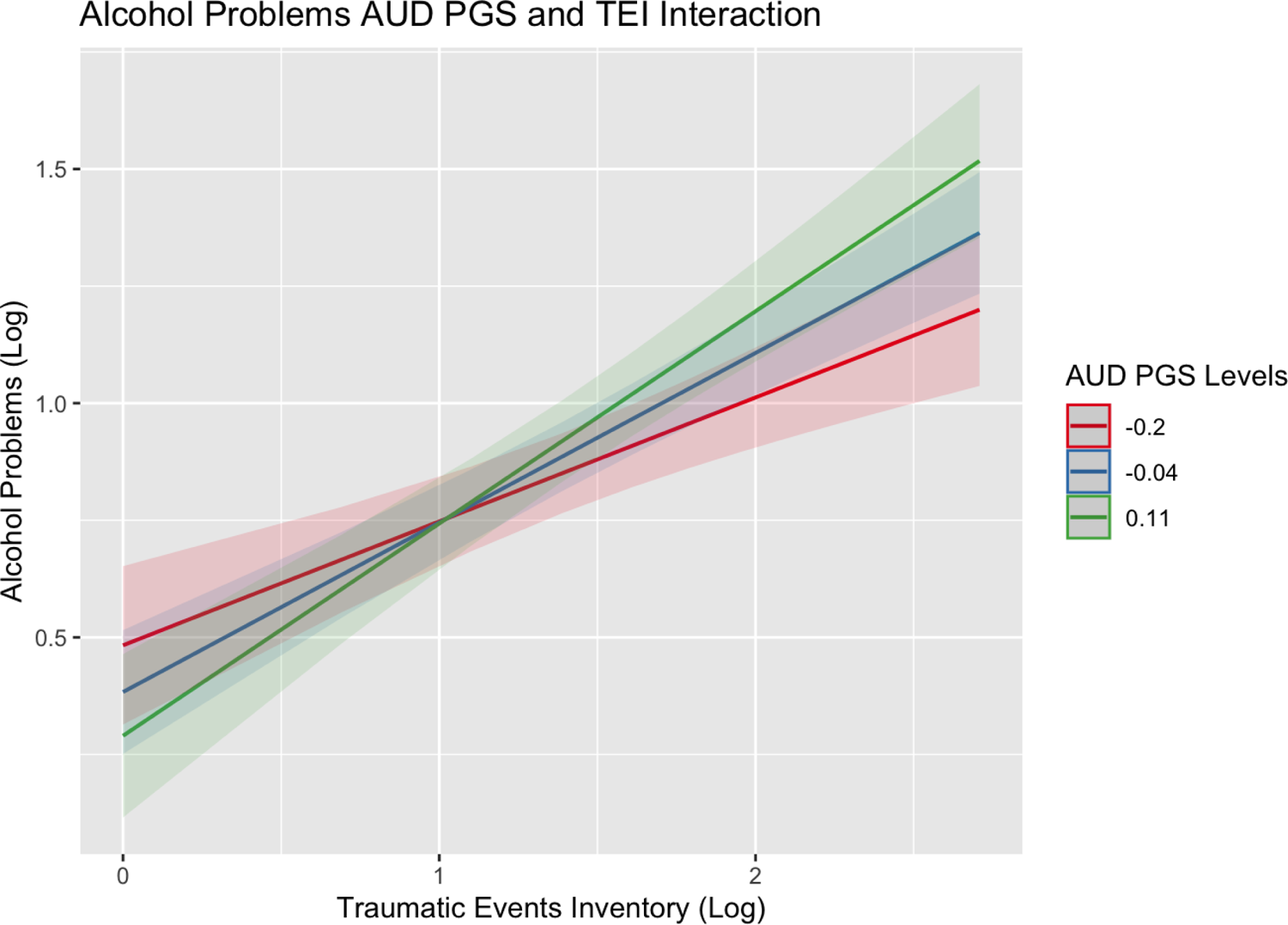
Alcohol problems interaction plot for PGS_AUD_ and TEI Figure showing the effects of lifetime trauma exposure on levels of alcohol problems among individuals with a PGS_AUD_ score two standard deviations below the mean (red), at the mean (blue), or above the mean (green) of the sample.

**Figure 3.**
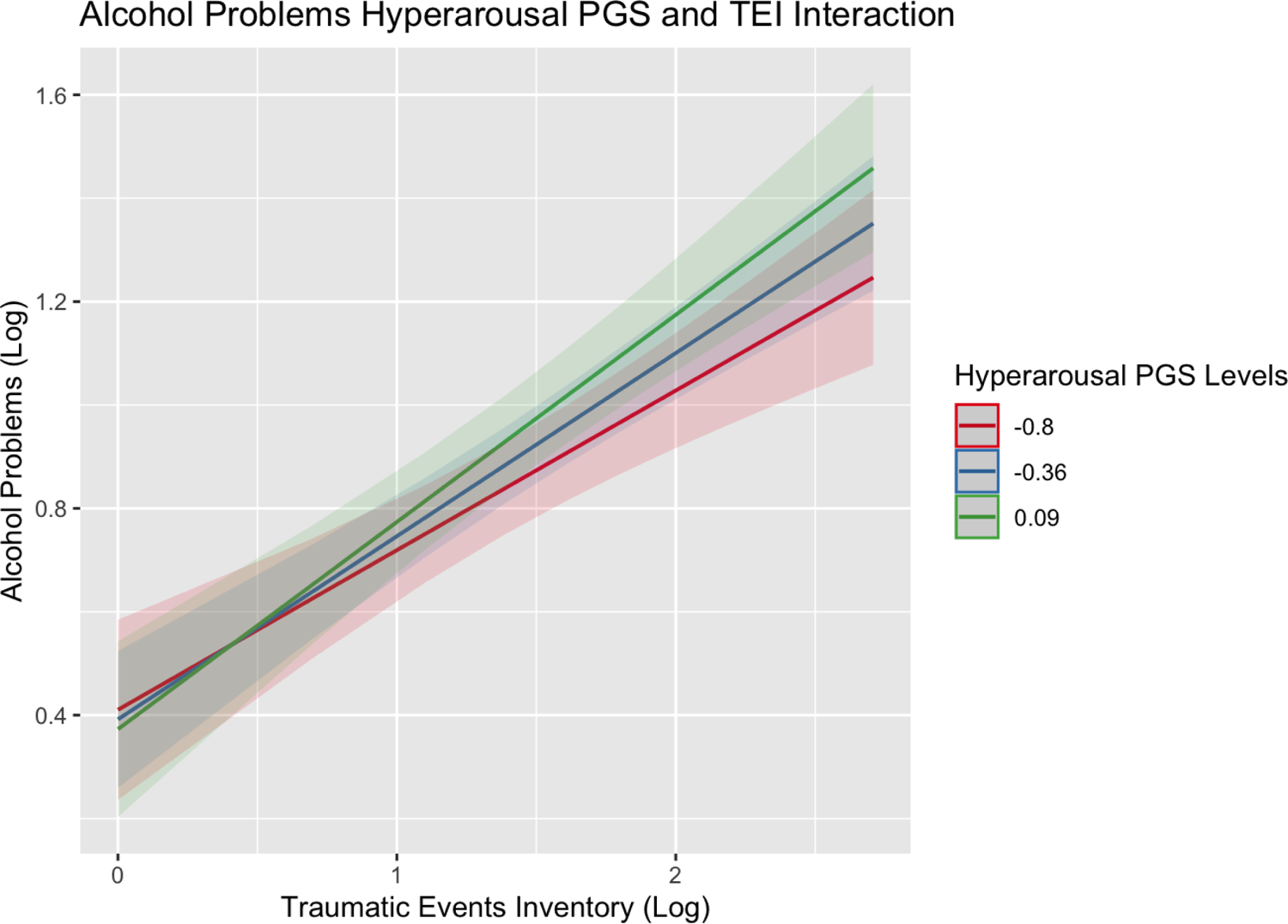
Alcohol problems interaction plot for PGS_HYPER_ and TEI Figure showing the effects of lifetime trauma exposure on levels of alcohol problems among individuals with a PGS_HYPER_ score two standard deviations below the mean (red), at the mean (blue), or above the mean (green) of the sample.

## DISCUSSION

The current study aimed to characterize the interplay of polygenic effects and traumatic experiences. It additionally fills a gap in the literature to provide evidence of this interplay in a population of Black people, given that most GWASs focus on outcomes in people of European descent due to increased sample sizes in that population. This study was able to leverage more recent efforts to increase sample sizes in individuals of African ancestry and utilize innovative tools that allow for the application of GWAS findings across ancestries using LD. We found support for several of our hypotheses. First, as expected, both a history of maltreatment during childhood maltreatment and lifetime trauma exposure were associated with alcohol consumption and problematic use during adulthood. The evidence for our second hypothesis was more nuanced. Those with predisposing genetic liability for alcohol use, AUD, and PTSD showed more problematic drinking, but not necessarily more alcohol consumption, unless the relationship with prior traumatic experience was considered.

Our models focused on individual PGS associations in the context of important demographics (e.g., age, sex, education levels) but not accounting for prior traumatic experiences (see supplemental information) demonstrated significant associations between PGSs for alcohol consumption, AUD, the PTSD sub-domains with alcohol problems. This illustrates the genetic risk for alcohol problems compared to alcohol consumption. Moreover, this differed from the zero order correlations, indicating that one or more of the demographic covariates was obscuring the relationship between polygenic influences of PTSD and alcohol behaviors on alcohol problems. However, our study also highlights the importance of contextual life factors, in this case trauma history, when attempting to interpret polygenic scores. We saw evidence of confounding effects among PGS effects in our graduated models. Specifically, once prior trauma was accounted for in models, the effect sizes of the PGSs effects on alcohol problems decreased, where only two PTSD subdomain PGSs (PGS_REEX_ and PGS_AVOID_) remained significantly associated with alcohol problems. This suggests that the effects of trauma and these polygenic scores are partially confounded, or that both trauma and polygenic scores explain some of the same variance in alcohol problems. It is important to note that the genetic risk for PTSD re-experiencing and avoidance symptoms (i.e., PGS_REEX_ and PGS_AVOID_) and prior traumatic experiences explain unique variance in alcohol problems. This may point to genetic influences on a person’s response to maltreatment and trauma that are unique within the re-experiencing and avoidance domains that then contribute to problematic alcohol use. This pattern of results was very different compared to alcohol consumption.

Our results and prior literature on alcohol consumption suggest that experiencing more childhood maltreatment (45, 46) and lifetime traumatic experiences (47-49) are significantly related to increased alcohol consumption as an adult. In general, polygenic scores for alcohol and PTSD were not associated with alcohol consumption with one exception. Individuals with higher polygenic scores for AUD saw a more drastic increase in their alcohol consumption when they had also experienced more traumatic events within their lifetime. These analyses indicate that understanding a person’s prior trauma and maltreatment history may be more relevant for a person’s level of alcohol consumption than understanding their genetic predisposition for alcohol or PTSD phenotypes at this time.

The current findings should be interpreted given several limitations. Given that current employment status is correlated with problematic alcohol use but not associated in the regression models that include trauma and polygenic scores, employment status is confounded with one of the other variables in the model. Further analyses and quasi-experimental hypotheses are needed to explore mechanisms that might explain this confound in a larger and more representative sample of the diversity of socioeconomics among African Americans (50). For instance, severe PTSD can impact a person’s ability to work, and in some cases where that occurs, individuals may qualify for social security benefits or disability benefits from the Veterans Affairs (51). If PTSD severity or disability status (which were not examined in the current study) are impacting employment status, then trauma and/or PTSD PGSs may be the confounding factors depending on the extent to which they are related to PTSD severity and/or disability benefits. Lastly, these analyses are exploratory and in need of further replication. PGS effects for trauma were correlated highly with each other and uncorrelated with genetic effects on alcohol consumption and problems. We focus on the estimates and standard errors for each model rather than interpreting solely based on the p-values as any attempt to correct for multiple testing would likely have been an overcorrection given the lack of independence across these PGSs.

Overall, the current findings suggest that some individuals may be more likely to experience problematic drinking due to genetic influences, even when levels of trauma are accounted for. Those who have higher genetic predisposition towards the reexperiencing or avoidance PTSD subdomain symptoms may experience more problematic drinking compared to those with a lower genetic predisposition even if they experience the same number of traumatic events. Additionally, some individuals may experience more problematic drinking if they have a higher genetic predisposition towards AUD or hyperarousal PTSD symptoms if they experience more traumatic events compared to those with lower genetic predisposition or those who experience fewer traumatic events. These genetic influences may be contributing to an individual’s response to trauma and maltreatment, which in turn leads to more problematic drinking. However, the effect sizes of these genetic influences remain small, and prevention of maltreatment and traumatic events remains one of the strongest ways to reduce problematic drinking behaviors as a response to trauma. While prevention is not always possible, providing the appropriate support systems and resources to individuals who experience maltreatment and trauma also remains crucial to lessen the environmental and genetic impacts of trauma on alcohol use.

## Data Availability

All data produced in the present study are available upon reasonable request to the authors.

## Notes

Primary Funding: This work was supported by grants from the National Institute of Health: R01DA042742, K23AT009713, and MH071537.

### Competing Interest Statement

The authors have declared no competing interest.

### Funding Statement

This work was supported by grants from the National Institute of Health: R01DA042742, K23AT009713, and MH071537.

### Author Declarations

Ethics committee/IRB of Emory University gave ethical approval for this work.

