## Supplemental Information for "Polygenic and *Polygene x Trauma* Contributions to Alcohol Use and Problems Among Black Americans"

### Supplementary Information

### Methods

#### Summary Statistics

**Alcohol Consumption (AUDIT-C) and Alcohol Use Disorder (AUD).** Kranzler and colleagues (2019) conducted a GWAS for both AUDIT-C and AUD. The ancestral groups that Kranzler et al. (2019) included in their multi-ancestral GWAS were: European Americans, African Americans, Latino Americans, East Asian Americans, and South Asian Americans. AUDIT-C was measured through the total scores of the first three items of the AUDIT (Kranzler et al., 2019). AUD was assessed through an AUD diagnosis using the International Classification of Disorders (ICD) (Kranzler et al., 2019). The ICD codes that Kranzler and colleagues (2019) looked at were: for the ICD-9, 303.X, which assesses dependence, and 305-305.03, which assessed abuse; for the ICD-10, F10.1 and F10.2, which assess dependence and abuse respectively. Participants received an AUD diagnosis if, from the years 2000 to 2008, they had at least one outpatient or two outpatient alcohol-associated ICD-9 or ICD-10 codes (Kranzler et al., 2019).

**PTSD-Hyperarousal, PTSD-Avoidance, PTSD-Reexperiencing, and Total PCL Score.** Stein and colleagues (2021) conducted a GWAS that examined PTSD and three subdomains of PTSD symptoms. The subdomains they examined were hyperarousal, avoidance, and reexperiencing. There were several steps Stein et al. (2021) conducted to classify a participant as a PTSD case. First, Stein and colleagues (2021) identified significant predictors of PTSD through the electronic health record of the sample. Next, Stein et al. (2021), using Lasso-regression, created a predicted probability score of having PTSD for every individual in the sample - overall, participants were classified in one of three ways: likely PTSD, possible PTSD, and no PTSD. Finally, they used a 0.70 probability threshold to determine which participants were classified as cases and which were classified as controls (Stein et al., 2021).

Participants were also given the DSM-IV version of the PTSD Symptom Checklist (PCL), which includes items that inquire about the symptoms of the three aforementioned subdomains of PTSD symptoms (Stein et al., 2021). The hyperarousal domain was assessed through 5 items in the PCL, and participants answered using a 5-point Likert scale (Stein et al., 2021). Similarly, the avoidance domain was assessed through 7 items in the PCL, and again, participants responded via a 5-point Likert scale (Stein et al., 2021). Stein and colleagues (2021) also included the total PCL score in their association analyses.

For their trans-ancestral GWAS, they collected information from 214,408 European Americans and 51,036 African Americans (Stein et al., 2021). We only included the African American sample to create our PGSs - there were 11,920 individuals who were identified as likely having PTSD and 39,116 controls (Stein et al., 2021).

<u>**PTSD Reexperiencing:**</u> Gelernter and colleagues (2019) conducted a GWAS that focused on the reexperiencing subdomain of PTSD symptoms. Reexperiencing was assessed through five items of the PCL, and scores could range from 5 to 25 (Gelernter et al., 2019). Gelernter et al. (2019) identified 146,660 European Americans and 19,983 African Americans from the MVP to include in their analyses. We only included the data for African Americans in our study.

Table S1. Effects of Traumas and PGSs Separately on Alcohol Consumption

| **Model** | **1. TEI** | **2. CTQ** | **3. PGS$_{AUDIT-C}$** | **4. PGS$_{AUD}$** | **5. PGS$_{REEX}$** | **6. PGS$_{HYPER}$** | **7. PGS$_{AVOID}$** | **8. PGS$_{PCL}$** |
|---|---|---|---|---|---|---|---|---|
| | **est (se)** | **est (se)** | **est (se)** | **est (se)** | **est (se)** | **est (se)** | **est (se)** | **est (se)** |
| **Predictor** | **0.28 (0.02)** | **0.17 (0.02)** | 0.01 (0.02) | 0.03 (0.02) | 0.04 (0.03) | 0.03 (0.02) | 0.02 (0.03) | 0.02 (0.03) |
| **Sex** | **-0.22 (0.02)** | **-0.29 (0.02)** | **-0.26 (0.02)** | **-0.26 (0.02)** | **-0.26 (0.02)** | **-0.26 (0.02)** | **-0.26 (0.02)** | **-0.26 (0.02)** |
| **Age** | **0.18 (0.02)** | **0.22 (0.02)** | **0.20 (0.02)** | **0.20 (0.02)** | **0.20 (0.02)** | **0.20 (0.02)** | **0.20 (0.02)** | **0.20 (0.02)** |
| **Education** | **-0.06 (0.02)** | -0.03 (0.02) | -0.04 (0.02) | -0.04 (0.02) | -0.04 (0.02) | -0.04 (0.02) | -0.04 (0.02) | -0.04 (0.02) |
| **Employment** | -0.02 (0.02) | -0.02 (0.02) | -0.03 (0.02) | -0.03 (0.02) | -0.03 (0.02) | -0.03 (0.02) | -0.03 (0.02) | -0.03 (0.02) |
| **Income** | -0.01 (0.02) | 0.01 (0.02) | -0.01 (0.02) | -0.01 (0.02) | -0.01 (0.02) | -0.01 (0.02) | -0.01 (0.02) | -0.01 (0.02) |
| **PC1** | -0.02 (0.02) | -0.01 (0.02) | 0.00 (0.02) | 0.00 (0.02) | 0.03 (0.03) | 0.01 (0.02) | 0.01 (0.03) | 0.02 (0.03) |
| **PC2** | 0.04 (0.02) | **0.04 (0.02)** | 0.04 (0.02) | 0.04 (0.02) | 0.04 (0.02) | 0.04 (0.02) | 0.04 (0.02) | 0.04 (0.02) |
| **PC3** | 0.02 (0.02) | 0.02 (0.02) | 0.02 (0.02) | 0.02 (0.02) | 0.03 (0.02) | 0.03 (0.02) | 0.03 (0.02) | 0.03 (0.02) |
| **PC4** | 0.01 (0.02) | 0.01 (0.02) | 0.01 (0.02) | 0.01 (0.02) | 0.01 (0.02) | 0.01 (0.02) | 0.01 (0.02) | 0.01 (0.02) |
| **PC5** | 0.03 (0.02) | 0.02 (0.02) | 0.03 (0.02) | 0.03 (0.02) | 0.02 (0.02) | 0.03 (0.02) | 0.02 (0.02) | 0.02 (0.02) |

**Note.** Bolded values met a $p < .05$ threshold; TEI = Trauma Experiences Inventory; CTQ = Childhood Trauma Questionnaire; AUDIT-C = AUDIT consumption subscore; AUD = Alcohol Use Disorder; REEX= Re-experiencing PTSD subdomain; HYPER= Hyperarousal PTSD subdomain; AVOID= Avoidance PTSD subdomain; PCL = PTSD Checklist Total score; PC = Principal Component.

Table S2. Effects of Traumas and PGSs Separately on Alcohol Problems

| **Model** | **1. TEI** | **2. CTQ** | **3. $\text{PGS}_{\text{AUDIT-C}}$** | **4. $\text{PGS}_{\text{AUD}}$** | **5. $\text{PGS}_{\text{REEX}}$** | **6. $\text{PGS}_{\text{HYPER}}$** | **7. $\text{PGS}_{\text{AVOID}}$** | **8. $\text{PGS}_{\text{PCL}}$** |
|---|---|---|---|---|---|---|---|---|
| | **est (se)** | **est (se)** | **est (se)** | **est (se)** | **est (se)** | **est (se)** | **est (se)** | **est (se)** |
| **Predictor** | **0.27 (0.02)** | **0.25 (0.02)** | **0.05 (0.02)** | **0.05 (0.02)** | **0.10 (0.03)** | **0.05 (0.02)** | **0.08 (0.03)** | **0.06 (0.03)** |
| **Sex** | **-0.21 (0.02)** | **-0.29 (0.02)** | **-0.25 (0.02)** | **-0.25 (0.02)** | **-0.25 (0.02)** | **-0.25 (0.02)** | **-0.25 (0.02)** | **-0.25 (0.02)** |
| **Age** | **0.09 (0.02)** | **0.13 (0.02)** | **0.12 (0.02)** | **0.12 (0.02)** | **0.12 (0.02)** | **0.12 (0.02)** | **0.12 (0.02)** | **0.12 (0.02)** |
| **Education** | **-0.14 (0.02)** | **-0.10 (0.02)** | **-0.11 (0.02)** | **-0.11 (0.02)** | **-0.11 (0.02)** | **-0.11 (0.02)** | **-0.11 (0.02)** | **-0.11 (0.02)** |
| **Employment** | -0.02 (0.02) | -0.02 (0.02) | -0.04 (0.02) | -0.04 (0.02) | -0.04 (0.02) | -0.03 (0.02) | -0.04 (0.02) | -0.04 (0.02) |
| **Income** | **-0.08 (0.02)** | **-0.06 (0.02)** | **-0.08 (0.02)** | **-0.08 (0.02)** | **-0.08 (0.02)** | **-0.08 (0.02)** | **-0.08 (0.02)** | **-0.08 (0.02)** |
| **PC1** | -0.01 (0.02) | 0.00(0.02) | 0.00 (0.02) | 0.01 (0.02) | **0.07 (0.03)** | 0.03 (0.02) | 0.06 (0.03) | 0.04 (0.03) |
| **PC2** | 0.02 (0.02) | 0.02 (0.02) | 0.02 (0.02) | 0.02 (0.02) | 0.02 (0.02) | 0.02 (0.02) | 0.02 (0.02) | 0.02 (0.02) |
| **PC3** | 0.03 (0.02) | 0.03 (0.02) | 0.03 (0.02) | 0.02 (0.02) | 0.04 (0.02) | 0.03 (0.02) | 0.04 (0.02) | 0.03 (0.02) |
| **PC4** | 0.04 (0.02) | 0.03 (0.02) | 0.04 (0.02) | 0.04 (0.02) | **0.05 (0.02)** | 0.04 (0.02) | 0.04 (0.02) | 0.04 (0.02) |
| **PC5** | 0.00 (0.02) | -0.00 (0.02) | 0.00 (0.02) | 0.00 (0.02) | 0.00 (0.02) | 0.00 (0.02) | -0.00 (0.02) | 0.00 (0.02) |

Note. Bolded values met a $p < .05$ threshold; TEI = Trauma Experiences Inventory; CTQ = Childhood Trauma Questionnaire AUDIT-C = AUDIT consumption subscore; AUD = Alcohol Use Disorder; REEX= Re-experiencing; PTSD subdomain; HYPER= Hyperarousal PTSD subdomain; AVOID= Avoidance PTSD subdomain; PCL = PTSD Checklist Total score; PC = Principal Component.

Table S3. Interaction Effects of Traumas and PGSs Separately on Alcohol Consumption

| **Model** | **3. $PGS_{AUDIT-C}$** | **4. $PGS_{AUD}$** | **5. $PGS_{REEX}$** | **6. $PGS_{HYPER}$** | **7. $PGS_{AVOID}$** | **8. $PGS_{PCL}$** |
|---|---|---|---|---|---|---|
| | **est (se)** | **est (se)** | **est (se)** | **est (se)** | **est (se)** | **est (se)** |
| **PRSCS PGS** | 0.09 (0.21) | 0.21 (0.21) | 0.12 (0.194) | 0.23 (0.20) | 0.21 (0.20) | 0.16 (0.20) |
| **TEI_LOG** | **0.26 (0.03)** | **0.26 (0.02)** | **0.27 (0.037)** | **0.26 (0.03)** | **0.26 (0.04)** | **0.26 (0.04)** |
| **CTQ_LOG** | **0.06 (0.03)** | **0.05 (0.02)** | 0.04 (0.036) | 0.04 (0.03) | 0.03 (0.04) | 0.04 (0.04) |
| **SEX** | **-0.23 (0.02)** | **-0.23 (0.02)** | **-0.24 (0.021)** | **-0.24 (0.02)** | **-0.23 (0.02)** | **-0.24 (0.02)** |
| **AGE** | **0.19 (0.02)** | **0.19 (0.02)** | **0.19 (0.02)** | **0.19 (0.02)** | **0.19 (0.02)** | **0.19 (0.02)** |
| **EDUCATION** | **-0.06 (0.02)** | **-0.06 (0.02)** | **-0.06 (0.02)** | **-0.06 (0.02)** | **-0.06 (0.02)** | **-0.06 (0.02)** |
| **EMPLOYMENT** | -0.02 (0.02) | -0.02 (0.02) | -0.02 (0.021) | -0.01 (0.02) | -0.01 (0.02) | -0.01 (0.02) |
| **INCOME2** | -0.01 (0.02) | -0.01 (0.02) | -0.01 (0.021) | -0.01 (0.02) | -0.01 (0.02) | -0.01 (0.02) |
| **PC1** | -0.02 (0.02) | -0.02 (0.02) | -0.01 (0.027) | -0.01 (0.02) | -0.01 (0.03) | -0.01 (0.03) |
| **PC2** | 0.04 (0.02) | 0.04 (0.02) | 0.04 (0.02) | 0.04 (0.02) | 0.04 (0.02) | 0.04 (0.02) |
| **PC3** | 0.02 (0.02) | 0.02 (0.02) | 0.02 (0.02) | 0.03 (0.02) | 0.02 (0.02) | 0.02 (0.02) |
| **PC4** | 0.01 (0.02) | 0.01 (0.02) | 0.01 (0.02) | 0.01 (0.02) | 0.01 (0.02) | 0.01 (0.02) |
| **PC5** | 0.03 (0.02) | 0.03 (0.02) | 0.03 (0.02) | 0.03 (0.02) | 0.03 (0.02) | 0.03 (0.02) |
| **TEIxPGS** | 0.05 (0.06) | **0.12 (0.05)** | 0.04 (0.063) | 0.05 (0.06) | 0.02 (0.06) | 0.03 (0.06) |
| **CTQxPGS** | -0.13 (0.22) | -0.31 (0.22) | -0.14 (0.212) | -0.26 (0.22) | -0.22 (0.22) | -0.17 (0.22) |

Note. Bolded values met a $p < .05$ threshold; TEI = Trauma Experiences Inventory; CTQ = Childhood Trauma Questionnaire AUDIT-C = AUDIT consumption subscore; AUD = Alcohol Use Disorder; REEX= Re-experiencing; PTSD subdomain; HYPER= Hyperarousal PTSD subdomain; AVOID= Avoidance PTSD subdomain; PCL = PTSD Checklist Total score; PC = Principal Component.

Table S4. Interaction Effects of Traumas and PGSs Separately on Alcohol Problems

| **Model** | **3. $PGS_{AUDIT-C}$** | **4. $PGS_{AUD}$** | **5. $PGS_{REEX}$** | **6. $PGS_{HYPER}$** | **7. $PGS_{AVOID}$** | **8. $PGS_{PCL}$** |
|---|---|---|---|---|---|---|
| | **est (se)** | **est (se)** | **est (se)** | **est (se)** | **est (se)** | **est (se)** |
| **PGS** | 0.27 (0.21) | 0.26 (0.21) | 0.03 (0.20) | 0.36 (0.20) | 0.23 (0.20) | 0.20 (0.20) |
| **TEI_LOG** | **0.22 (0.03)** | **0.22 (0.02)** | **0.24 (0.04)** | **0.23 (0.03)** | **0.26 (0.04)** | **0.24 (0.04)** |
| **CTQ_LOG** | **0.14 (0.03)** | **0.15 (0.02)** | **0.15 (0.04)** | **0.13 (0.03)** | **0.12 (0.04)** | **0.13 (0.04)** |
| **SEX** | **-0.24 (0.02)** | **-0.24 (0.02)** | **-0.24 (0.02)** | **-0.25 (0.02)** | **-0.25 (0.02)** | **-0.25 (0.02)** |
| **AGE** | **0.11 (0.02)** | **0.11 (0.02)** | **0.11 (0.02)** | **0.11 (0.02)** | **0.11 (0.02)** | **0.11 (0.02)** |
| **EDUCATION** | **-0.12 (0.02)** | **-0.12 (0.02)** | **-0.12 (0.02)** | **-0.12 (0.02)** | **-0.12 (0.02)** | **-0.12 (0.02)** |
| **EMPLOYMENT** | -0.01 (0.02) | -0.01 (0.02) | -0.01 (0.02) | -0.01 (0.02) | -0.01 (0.02) | -0.01 (0.02) |
| **INCOME2** | **-0.07 (0.02)** | **-0.07 (0.02)** | **-0.07 (0.02)** | **-0.07 (0.02)** | **-0.07 (0.02)** | **-0.07 (0.02)** |
| **PC1** | -0.01 (0.02) | -0.01 (0.02) | 0.04 (0.03) | 0.01 (0.02) | 0.04 (0.03) | 0.02 (0.03) |
| **PC2** | 0.02 (0.02) | 0.02 (0.02) | 0.02 (0.02) | 0.02 (0.02) | 0.02 (0.02) | 0.02 (0.02) |
| **PC3** | 0.03 (0.02) | 0.03 (0.02) | 0.03 (0.02) | 0.03 (0.02) | 0.04 (0.02) | 0.03 (0.02) |
| **PC4** | 0.03 (0.02) | 0.03 (0.02) | 0.04 (0.02) | 0.04 (0.02) | 0.03 (0.02) | 0.04 (0.02) |
| **PC5** | 0.00 (0.02) | 0.00 (0.02) | 0.00 (0.02) | 0.00 (0.02) | 0.00 (0.02) | 0.00 (0.02) |
| **TEIxPGS** | 0.09 (0.06) | **0.17 (0.05)** | 0.10 (0.06) | **0.11 (0.06)** | 0.12 (0.06) | 0.09 (0.06) |
| **CTQxPGS** | -0.32 (0.22) | -0.38 (0.22) | -0.03 (0.21) | -0.42 (0.22) | -0.26 (0.22) | -0.23 (0.22) |

Note. Bolded values met a $p < .05$ threshold; TEI = Trauma Experiences Inventory; CTQ = Childhood Trauma Questionnaire AUDIT-C = AUDIT consumption subscore; AUD = Alcohol Use Disorder; REEX= Re-experiencing; PTSD subdomain; HYPER= Hyperarousal PTSD subdomain; AVOID= Avoidance PTSD subdomain; PCL = PTSD Checklist Total score; PC = Principal Component.
